# Chemoprophylaxis of COVID-19 with hydroxychloroquine - a study of health care workers attitude, adherence to regime and side effects

**DOI:** 10.1101/2020.06.11.20126359

**Authors:** Debajyoti Bhattacharyya, Neeraj Raizada, Bharathnag Nagappa, Arvind Tomar, Prateek Maurya, Ashok Chaudhary, Mini George, Deepty Katiyal, Srishti Rajora, Nikky Singh

## Abstract

**Background:** There are apprehensions amongst healthcare worker (HCWs) about COVID-19. The HCWs have been given hydroxychloroquine (HCQ) chemoprophylaxis for seven weeks as per Government of India guidelines.

**Objectives:** To assess the apprehensions amongst HCWs about COVID-19 and to document accessibility, adherence and side effects related to HCQ prophylaxis in HCWs.

**Methods:** A longitudinal follow up study was conducted in a tertiary care hospital. HCQ was given in the dose of 400 mg twice on day one, and then 400 mg weekly for seven weeks. 391 HCWs were interviewed using semi structured questionnaire.

**Results:** 62.2% HCWs expressed perceived danger posted by COVID-19 infection. Doctors (54%) showed least acceptance and paramedics (88%) showed highest acceptance to chemoprophylaxis. 17.5% participants developed at least one of the side effects to HCQ. Females and nursing profession were significantly associated with adverse effects. Common side effects were gastro-intestinal symptoms, headache and abnormal mood change. Most of these were mild, not requiring any intervention. Gender, professions and perceived threat of COVID-19 were significantly associated with acceptance and adherence to HCQ prophylaxis.

**Conclusion:** Two third of HCWs had perceived danger due to COVID-19. Three fourth of the HCWs accepted chemoprophylaxis and four out of five who accepted had complete adherence to prophylaxis schedule. One out of five had developed at least one of side effects; however, most of these were mild not requiring any intervention.

## Introduction

The disease caused by SARS-CoV-2 is called COVID-19. It was first described in Wuhan province of China in December 2019,^1,2^ and since then it is spreading rapidly. Scientists are searching for specific anti-viral drug or vaccine against this disease.^3^ However there are conflicting results that a few drugs are showing promising results.^4,5^ Health care workers exposed to confirmed or suspected case of COVID-19 are at high risk of getting infected with COVID-19.^6^ In view of this, National task force for COVID-19 constituted by Indian Council of Medical Research (ICMR) under Government of India (GOI) has recommended hydroxychloroquine (HCQ) for adults in a dosage of 400 mg twice a day on first day followed by 400 mg once a week for 7 weeks for chemoprophylaxis for COVID-19 for all health care workers (HCWs) involved in the care of suspected or confirmed cases of COVID-19, and also for household contacts of laboratory confirmed cases.^7^

There are apprehensions amongst healthcare workers (HCWs) about COVID-19 as well as about taking HCQ^8–11^ as chemoprophylaxis because of possible side effects.^12-17^ There is currently limited experience related to administration of HCQ as chemoprophylaxis for COVID-19 as the general guidance is based on a small sample of data. This limitation may translate into its low acceptability in HCWs. There are a few clinical trials presently going on in the context of chemoprophylaxis with HCQ in COVID-19.^13^ Also, there have been a few reports of severe side-effects to HCQ in medical journals.^12-17^ These reports may lead to apprehensions in the target population while scaling up this chemoprophylaxis. Hence, it is felt that a study should explore and document experiences in large scale administration of HCQ chemoprophylaxis for COVID-19 in HCWs and also their attitude and confidence level before and after taking chemoprophylaxis. The well documented side effects of HCQ include gastrointestinal side effects like nausea, vomiting, gastritis, diarrhoea, allergic reactions like itching/rashes, eye problems like loss or decrease in accommodation, burning sensation, abnormal mood changes, giddiness and change in cardiac rhythm.^12-17^ However, adverse effects of HCQ during its application chemoprophylaxis for COVID-19 in the recommended dosages should also be explored. This study was conceived when HCQ chemoprophylaxis administration was initiated for being given to HCWs of our hospital for COVID-19 in response to Indian Council of Medical Research, Government of India guidance on the same.^7^

## Materials and Methods

### Study Design and setting

A longitudinal follow up study was conducted in a tertiary care hospital in Delhi. The hospital is a teaching institute with 400 beds, and is providing outpatient care for an average 200 patients a day. The hospital has a total of 689 employees which includes doctors, nurses, paramedic staffs and other supporting staffs.

### Study Procedure

Following the guidance issued by Government of India, recommending the use of HCQ for prophylaxis of COVID-19, HCQ chemoprophylaxis has been offered to all the healthcare staffs of the institute since 28^th^ March 2020. As a part of capacity building they were made aware on various aspects of HCQ prophylaxis, such as, available evidence, rationale, its side effects and asked to report if they developed any symptom for prompt management of the same. Subsequently, the current study was undertaken to assess different modalities associated with administration of this intervention. All the HCWs were invited to participate in the study, and consent was obtained from all of them who were willing to participate. A trained team of HCWs interviewed the participants with semi structured questionnaire, which included details about socio-demographic details, co-morbidities, perceived threat from COVID-19, source of knowledge regarding HCQ prophylaxis. All participants were followed up to assess the development of side effects (self-reported) and adherence to prophylaxis. All participants who developed the side effects were referred to physician for confirmation and further management of side effects. The data were captured using digital data entry form.

### Statistical methods

Data were extracted in Microsoft Excel and analysed in Statistical Package for Social Science version 22. Continuous variables were expressed with mean with standard deviation (SD). Categorical variables were expressed with proportions. Proportion of HCWs accepted prophylaxis, proportion with poor adherence, and who developed side effects were expressed as percentages with 95% Confidence interval (95% CI). Chi square test was used to test the statistical significance. p value less than 0.05 was considered statistically significant.

### Ethical Approval

This study protocol was approved by the institutional ethics committee.

## Results

Overall 391 health care workers with mean age of 34±8 years were included in the study. Around 229 (59%) of the participants were males and 147 (38%) were working as nurse. Around 86 (22%) were very scared, 84 (21%) were minimally scared and 99 (26%) did not express any opinion on perceived danger that was posed by COVID 19. Details on characteristics of study participants are given in table 1.

**Table 1:** Characteristics of study participants (n=391)

| Characteristics | number | % |
| --- | --- | --- |
| Age group |  |  |
| <30 years | 134 | 34.3 |
| 30 to 45 years | 220 | 56.3 |
| 45 to 60 years | 34 | 8.7 |
| >60 years | 3 | .8 |
| Gender |  |  |
| Male | 229 | 58.6 |
| Female | 162 | 41.4 |
| Profession |  |  |
| Administration | 31 | 7.9 |
| Doctor | 57 | 14.6 |
| Nursing | 147 | 37.6 |
| Paramedical | 124 | 31.7 |
| Others | 32 | 8.1 |
| Chronic diseases |  |  |
| Liver Disease | 5 | 1.2 |
| Cardiovascular disease | 8 | 1.9 |
| Kidney disease | 1 | 0.3 |
| Respiratory disease | 5 | 1.2 |
| Diabetes mellitus | 7 | 1.9 |
| No co-morbidities | 365 | 93.4 |
| *Perceived danger due to COVID 19* |  |  |
| Not scared at all | 49 | 12.5 |
| Minimally scared | 84 | 21.5 |
| Somehow scared | 73 | 18.7 |
| Very scared | 86 | 22.0 |
| No opinion | 99 | 26.3 |

In total, 297 (75.9%, 95% CI 71.7% - 80.2%) of study participants opted for hydroxychloroquine prophylaxis, among them 254 (85.5%, 95% CI 81.5% - 89.5%) had taken doses as per schedule. The acceptance of hydroxychloroquine prophylaxis was high among males (80%) and also among participants aged less than 30 years (78%) which gradually decreases as age increase, but it was not statistically significant. Among different professions, doctors (54%) showed least acceptance to HCQ prophylaxis and paramedics (88%) showed the highest acceptance and it was statistically significant (p value <0.0001). Details on factors associated with acceptance of hydroxychloroquine prophylaxis are given in table 2. Gender (p value 0.012), profession of study participants (0.003) and perceived danger due to COVID 19 (<0.0001) were significantly associated with compliance to prophylaxis (table 2). Reasons given for not taking prophylaxis are shown in table 3.

**Table 2:**
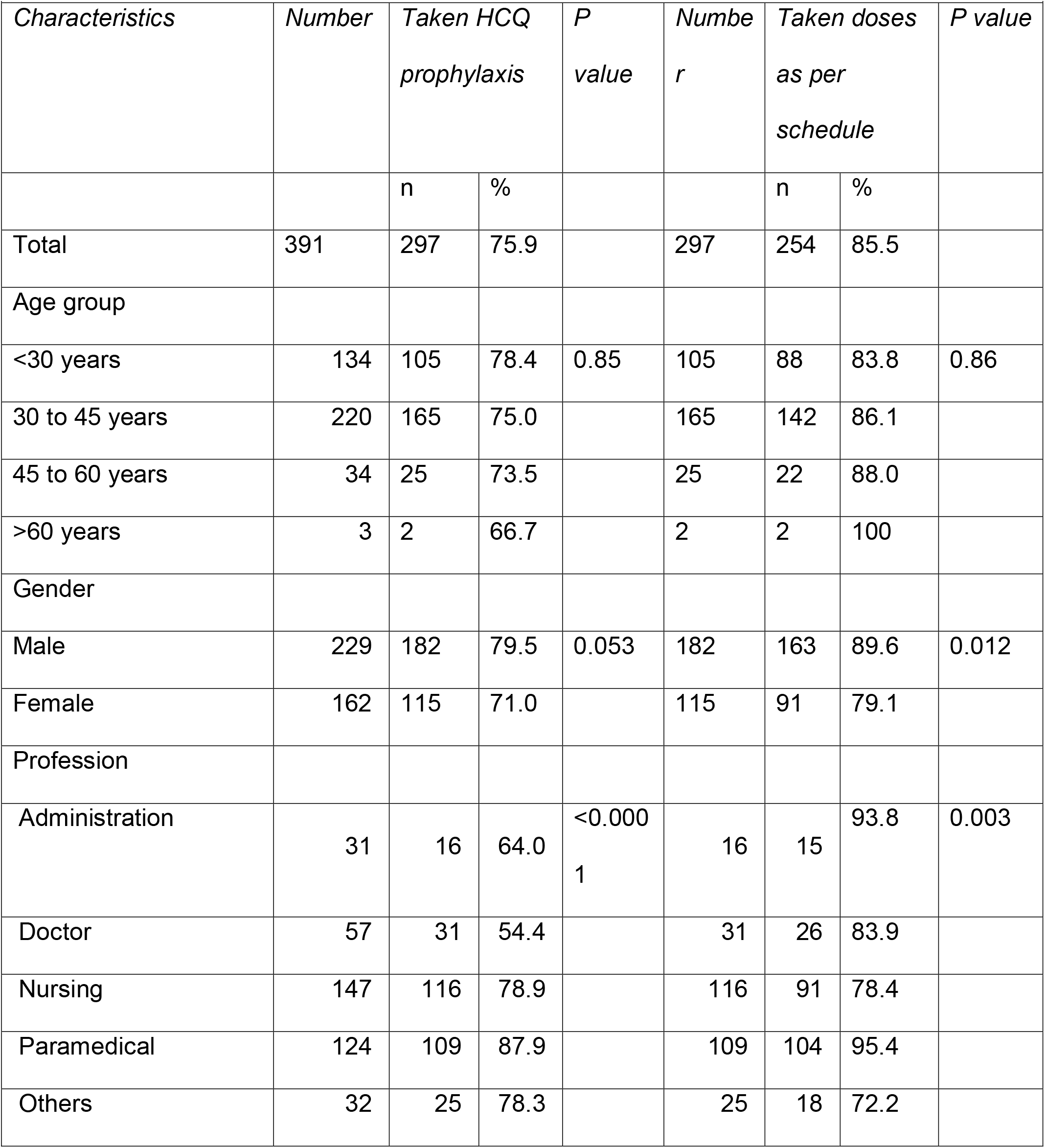
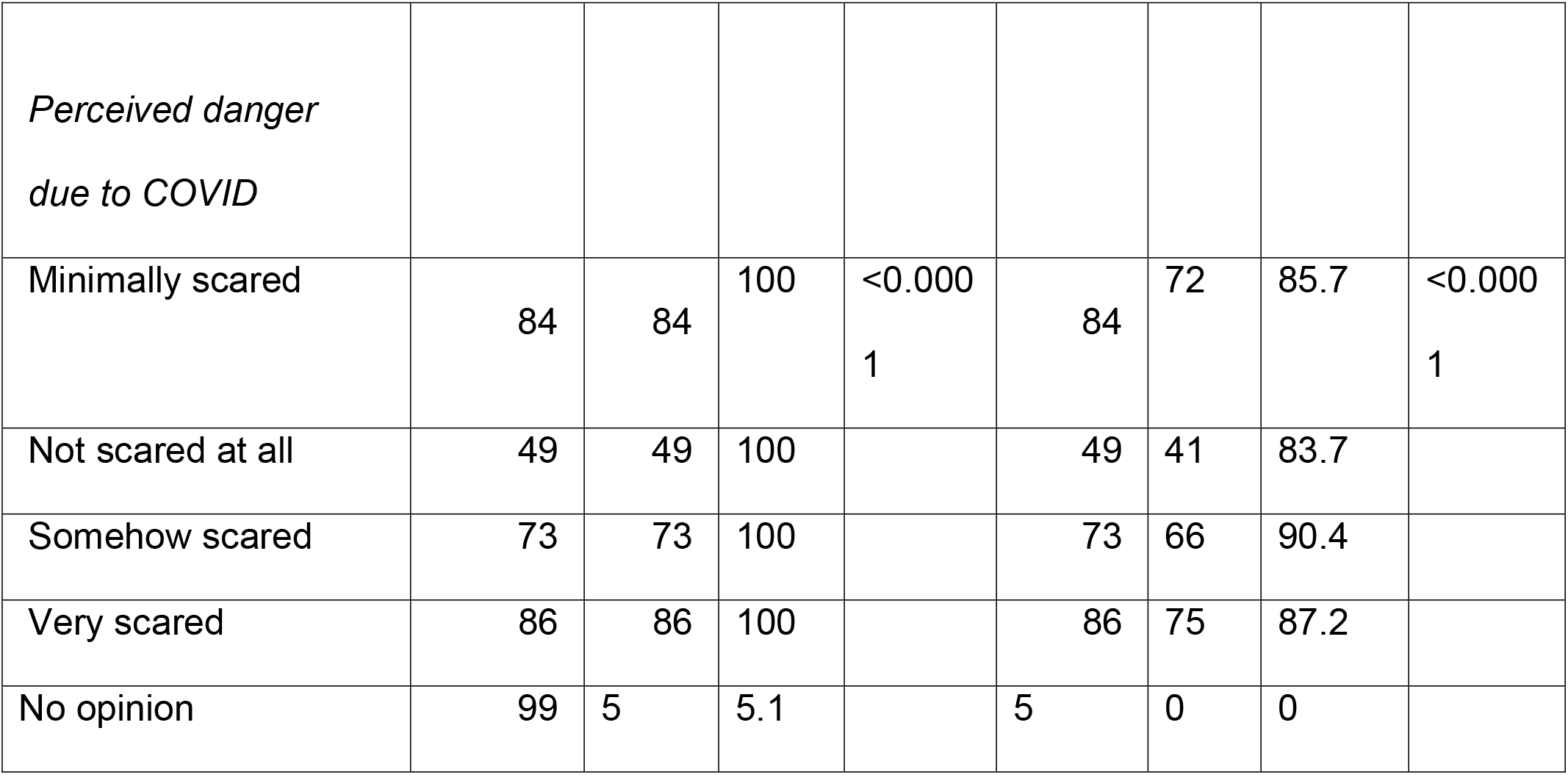
Factors associated with uptake and adherence to hydroxychloroquine prophylaxis for COVID-19 among health care workers

**Table 3.** Reasons given by health care workers for not taking hydroxychloroquine

| Characteristics | Frequency* | Proportion (%) |
| --- | --- | --- |
| Fear of adverse effect | 22 | 23.4 |
| On other medication | 12 | 12.8 |
| No clear evidence | 10 | 10.6 |
| Pregnant/lactating | 7 | 7.4 |
| Not exposed | 7 | 7.4 |
| Acute illness | 6 | 6.4 |
| Developed side effects | 3 | 3.2 |
| Not interested | 2 | 2.1 |
| Others | 10 | 10.6 |
| No valid reasons given | 23 | 24.5 |

\*categories are mutually inclusive

**Table 3a.**
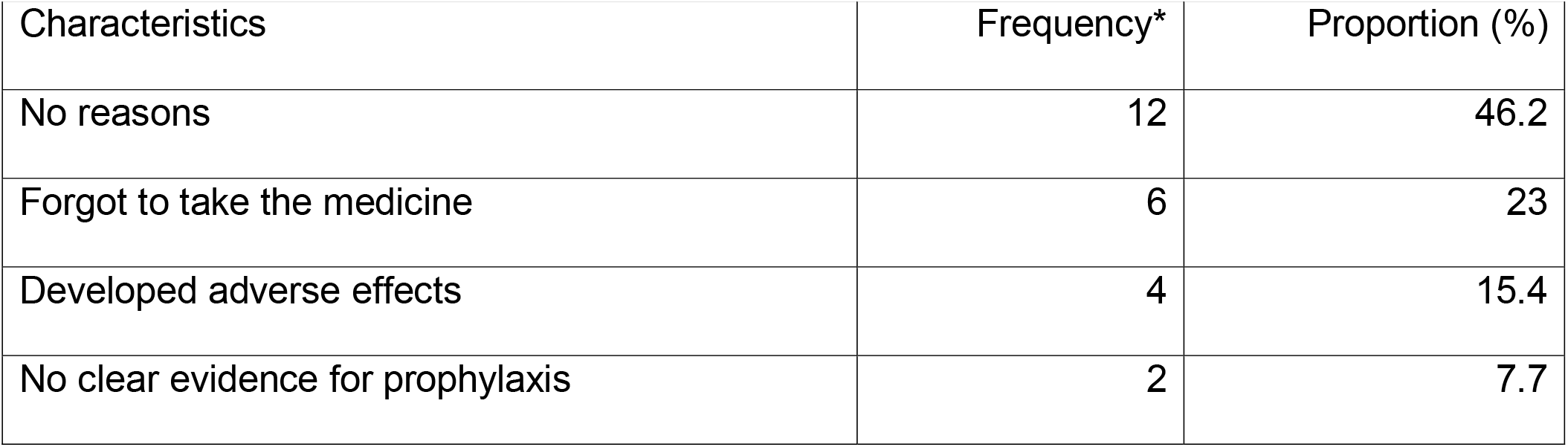
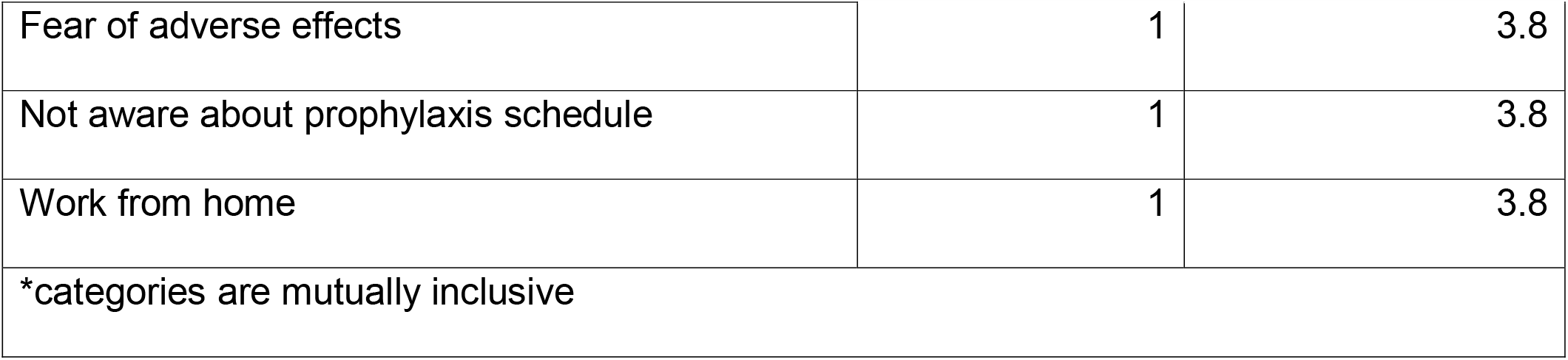
Reasons given by health care workers for discontinuing hydroxychloroquine prophylaxis (n=26)

Among the participants not accepted the prophylaxis, 22 (23.4%) did so for fear of side effects, 12 (12.8%) were taking other medications and 10 (10.6%) felt that there is no clear evidence supporting the efficacy of prophylaxis in preventing COVID 19. Among study participants who missed the dose or discontinued the prophylaxis, 6 (23%) forgot to take medication, 4 (15%) discontinued after developing adverse effects and 4 (8%) felt there is no clear evidence supporting the efficacy of prophylaxis in preventing COVID 19. Details on reasons for poor compliance to prophylaxis are given in table 3a.

Among the 297 participants taking hydroxychloroquine prophylaxis, 52 (17.5%, 95% CI 13.2% - 21.8%) developed adverse effects due to hydroxychloroquine prophylaxis. Of the 297 participants taking HCQ prophylaxis, 29 (9.8%) reported gastrointestinal adverse effects, 12 (4%) reported mild allergic reactions, 11 (3.7%) developed head ache and 9 (3%) participants reported abnormal mood change. Other side effects reported by study participants are shown in table 4. Females and nursing profession were significantly associated with adverse effects due to prophylaxis (table 5).

**Table 4:** Adverse effects due to hydroxychloroquine among health care workers (n=297)

|  |  |  |
| --- | --- | --- |
| Adverse effects | Frequency* | Proportion (%) |
| Gastro intestinal side effects (vomiting-10, diarrhoea -12, gastritis-7) | 29 | 9.8 |
| Allergic reaction | 12 | 4.0 |
| Headache | 11 | 3.7 |
| Abnormal mood change | 9 | 3.0 |
| Palpitation | 2 | 0.6 |
| Vertigo | 2 | 0.6 |
| Ocular symptoms | 1 | 0.3 |
| Hypotension | 1 | 0.3 |
| Heaviness in chest | 1 | 0.3 |
| Fever | 1 | 0.3 |
| Total | 52 | 17.5 |
| *categories are mutually inclusive |  |  |

**Table 5:**
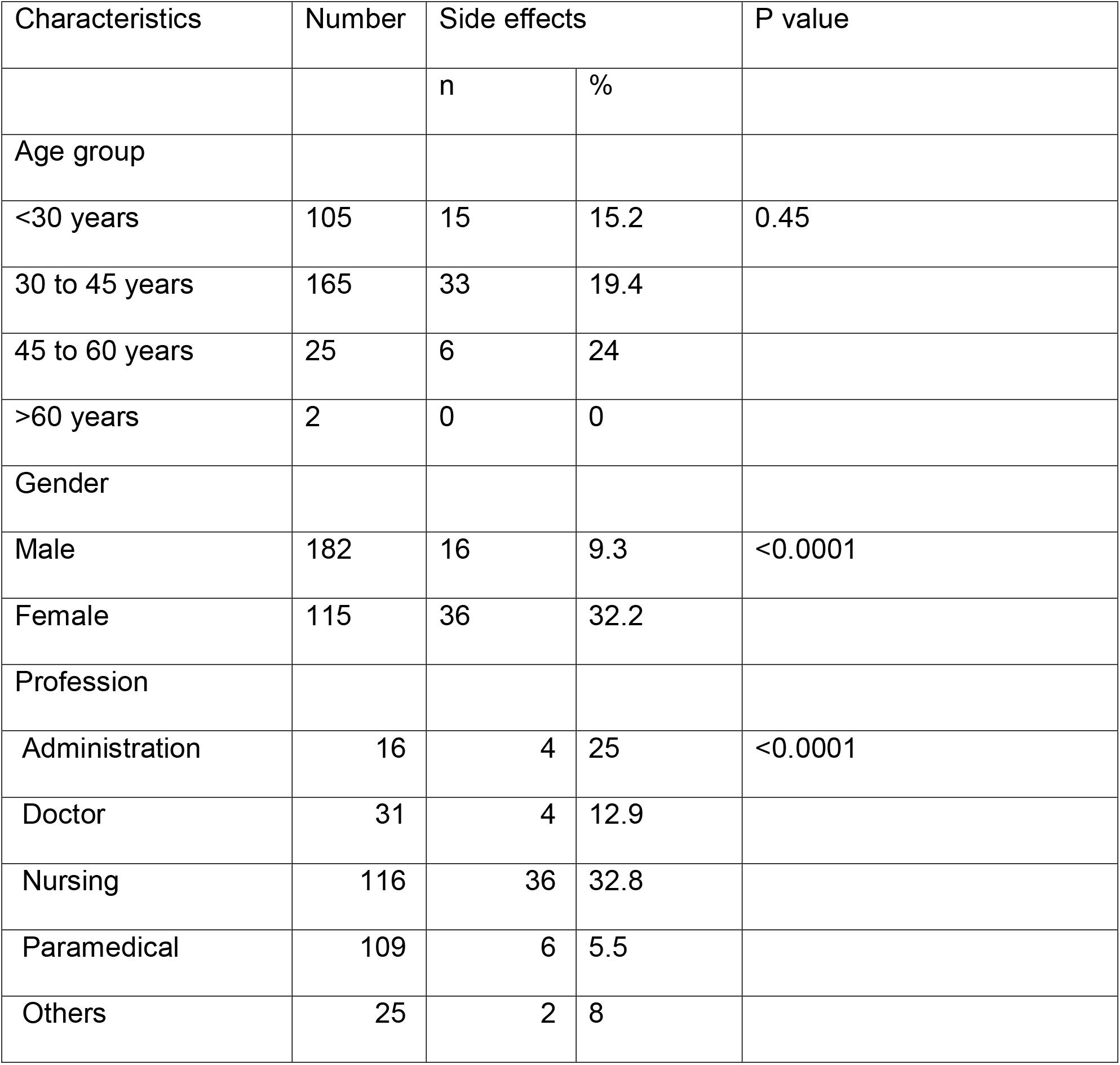
Factors associated with adverse effects due to hydroxychloroquine among health care workers (n=297)

| Characteristics | Number | Side effects |  | P value |
| --- | --- | --- | --- | --- |
|  |  | n | % |  |
| Age group |  |  |  |  |
| <30 years | 105 | 15 | 15.2 | 0.45 |
| 30 to 45 years | 165 | 33 | 19.4 |  |
| 45 to 60 years | 25 | 6 | 24 |  |
| >60 years | 2 | 0 | 0 |  |
| Gender |  |  |  |  |
| Male | 182 | 16 | 9.3 | <0.0001 |
| Female | 115 | 36 | 32.2 |  |
| Profession |  |  |  |  |
| Administration | 16 | 4 | 25 | <0.0001 |
| Doctor | 31 | 4 | 12.9 |  |
| Nursing | 116 | 36 | 32.8 |  |
| Paramedical | 109 | 6 | 5.5 |  |
| Others | 25 | 2 | 8 |  |

## Discussion

The present study was aimed to assess the acceptance, adherence and adverse effects related to HCQ chemoprophylaxis to prevent COVID-19 among health care workers. The present study is the first one to study the acceptance and adherence of HCQ prophylaxis for COVID-19. The study found that around 76% of the HCWs had accepted the prophylaxis and taken HCQ at least once as a part of prophylaxis. The finding is comparable with findings of the studies conducted among travellers to assess the acceptance to chloroquine/ mefloquine prophylaxis for malaria, which shows 52% to 89% acceptance rate.^18-21^ However, as the present study was conducted among health workers, we could expect a higher acceptance among our study population. As expressed by study participants, common reasons for not taking prophylaxis were fear of side effects, already on other medication and no existing clear evidence on effectiveness of chemoprophylaxis to prevent COVID-19.^22-30^ In particular fear of side effects could be attributed to several reports of side effects. These kinds of advices were not issued by reputed health organisations when chloroquine was considered as prophylaxis for malaria. Also, there is no clear evidence of effectiveness of prophylaxis against COVID-19 as it is against malaria. HCWs are well aware of these factors which lead to relatively lesser acceptance of prophylaxis. These findings further justified by significantly lower acceptance among doctors followed by nurse and higher acceptance among other supporting staff. Doctors were high likely to have knowledge on effectiveness and safety of prophylaxis. In the current era of evidence based medical practice, they are likely to reject prophylaxis due to even negligible uncertainty. Study also shows that the perceived threat due to COVID-19 is very less among doctors compared to other professions (details provided in supplementary table 1) and this may hamper the motivation to take chemoprophylaxis.

Among participants who had accepted the prophylaxis, 85% had shown complete adherence. Females found to have poor adherence compared to males and it is similar to findings reported in systematic review conducted by Ahluwalia et al to assess the factors affecting adherence of chloroquine chemoprophylaxis against malaria.^18^ The study also reported that as the severity of perceived threat due to COVID-19 increases, adherence level also increase. This may be due to effect of perceived threat on motivation to adhere to prophylaxis. Fear of getting infected by SARS-CoV-2, could have motivated them to strictly adhere to the prophylaxis. Study participants in nursing category had poor adherence, which may be due side effects (relatively higher proportion of nursing officers reported side effects, compared to others). Common reasons given for poor adherence to prophylaxis were forgetting to take drug and development of side effects and these findings were similar to the literature.^18^ Fear of side effects, no clear evidence and no clear knowledge about schedule of prophylaxis were other reasons for poor adherence. These findings suggest the need for awareness activities on safety, effectiveness and schedule of prophylaxis.

The study also found that around 18% of the study participants developed adverse effects related to prophylaxis. Common adverse effects were gastro intestinal related followed by allergic reactions and headache. Carme B et al reported that around 12% of people who had taken chloroquine prophylaxis to prevent malaria, had developed adverse effects which is comparable to our findings.^30^ Literature also shows that gastrointestinal adverse effects and head ache are the common adverse reactions of HCQ, which is similar to our study.^12-14,30^

Overall our study found that there is moderately good acceptance for the prophylaxis with good adherence. However still significant number of study participants had expressed fear of side effects and lack of clear evidence on effectiveness of prophylaxis, which should be addressed while giving mass advice. Also clear instructions need to be given to the people about when to stop prophylaxis if they develop adverse effects.

Strength of the present study are: 1) interview was conducted by trained HCWs in research, and data were entered in digital data entry form which would have minimised the error while collecting the data, and 2) all study participants were closely followed up regularly to assess development of any side effects by research team. The study has a few limitations like, 1) participation was entirely voluntary basis which may lead to selection bias, and 2) adherence to chemoprophylaxis was self-reported which may be slightly higher than actual adherence.

## Conclusion

Three fourth of the health care workers accepted chemoprophylaxis to prevent COVID 19 and four out of five accepted chemoprophylaxis had complete adherence to prophylaxis schedule. Gender, profession and perceived threats were significantly associated with acceptance and adherence. One out of five had developed at least one of the side effects with higher rate among females. However, side-effects were largely mild in nature, with less than 2% of the participants requiring to discontinue HCQ prophylaxis due to side effects.

## Data Availability

Raw data referred to in the manuscript is available at: https://drive.google.com/drive/folders/1PCazhITB8xjCT6fp_iHvjEHerDypmN9H

---

**Supplementary table 1:** Factors associated with perceived threat from COVID 19 among health care workers (n=391)

|  |  |  |  |  |  |  |
| --- | --- | --- | --- | --- | --- | --- |
| Characteristics | Expressed<br>no opinion | Not<br>scared | Minimally<br>scared | Somewhat<br>scared | Very<br>scared | P value |
| Gender |  |  |  |  |  |  |
| Females | 48<br>30.4% | 13<br>8.2% | 36<br>22.8% | 32<br>20.3% | 29<br>18.4% | 0.056 |
| Males | 51<br>22.3% | 36<br>15.7% | 44<br>19.2% | 41<br>17.9% | 57<br>24.9% |  |
| Age groups |  |  |  |  |  |  |
| <30 years | 31<br>23.1% | 19<br>14.2% | 32<br>23.9% | 24<br>17.9% | 27<br>20.1% | 0.66 |
| 30 to 45 years | 57<br>25.9% | 23<br>10.5% | 40<br>18.2% | 43<br>19.5% | 54<br>24.5% |  |
| 45 to 60 years | 10<br>29.4% | 7<br>20.6% | 6<br>17.6% | 6<br>17.6% | 5<br>14.7% |  |
| >60 years | 1 | 0 | 2 | 0 | 0 |  |
|  | 33.3% | 0.0% | 66.7% | 0.0% | 0.0% |  |
| Profession |  |  |  |  |  |  |
| Administration | 9 | 2 | 5 | 6 | 3 | <0.0001 |
|  | 36.0% | 8.0% | 20.0% | 24.0% | 12.0% |  |
| Doctor | 26 | 10 | 9 | 9 | 3 |  |
|  | 45.6% | 17.5% | 15.8% | 15.8% | 5.3% |  |
| Nursing | 33 | 16 | 35 | 34 | 26 |  |
|  | 22.4% | 10.9% | 23.8% | 23.1% | 17.7% |  |
| Others | 7 | 10 | 1 | 2 | 3 |  |
|  | 30.4% | 43.5% | 4.3% | 8.7% | 13.0% |  |
| Paramedical | 16 | 9 | 28 | 20 | 50 |  |
|  | 12.9% | 7.3% | 22.6% | 16.1% | 40.3% |  |

